# Comparisons of the risk of myopericarditis between COVID-19 patients and individuals receiving COVID-19 vaccines: a population-based study

**DOI:** 10.1101/2021.12.13.21267730

**Authors:** Oscar Hou In Chou, Jiandong Zhou, Teddy Tai Loy Lee, Thompson Kot, Sharen Lee, Abraham Ka Chung Wai, Wing Tak Wong, Qingpeng Zhang, Shuk Han Cheng, Tong Liu, Vassilios S Vassiliou, Bernard Man Yung Cheung, Gary Tse

## Abstract

**Background:** Both COVID-19 infection and COVID-19 vaccines have been associated with the development of myopericarditis. The objective of this study is to 1) analyze the rates of myopericarditis after COVID-19 infection and COVID-19 vaccination in Hong Kong and 2) compare to the background rates, and 3) compare the rates of myopericarditis after COVID-19 vaccination to those reported in other countries.

**Methods:** This was a population-based cohort study from Hong Kong, China. Patients with positive RT-PCR test for COVID-19 between 1^st^ January 2020 and 30^th^ June 2021 or individuals who received COVID-19 vaccination until 31^st^ August were included. The main exposures were COVID-19 positivity or COVID-19 vaccination. The primary outcome was myopericarditis.

**Results:** This study included 11441 COVID-19 patients from Hong Kong, of whom four suffered from myopericarditis (rate per million: 350; 95% confidence interval [CI]: 140-900). The rate was higher than the pre-COVID-19 background rate in 2020 (rate per million: 61, 95% CI: 55-67) with a rate ratio of 5.73 (95% CI: 2.23-14.73. Compared to background rates, the rate of myopericarditis among vaccinated subjects in Hong Kong was substantially lower (rate per million: 8.6; 95% CI: 6.4-11.6) with a rate ratio of 0.14 (95% CI: 0.10-0.19). The rates of myocarditis after vaccination in Hong Kong are comparable to those vaccinated in the United States, Israel, and the United Kingdom.

**Conclusions:** COVID-19 infection is associated with a higher rate of myopericarditis whereas COVID-19 vaccination is associated with a lower rate of myopericarditis compared to the background.

## Introduction

Since the beginning of the COVID-19 pandemic, cases of myocarditis and pericarditis related to the infection have been widely described.^1, 2^ However, recent studies have also reported possible associations between COVID-19 vaccines and the risk of myo-pericardial inflammation, raising concerns about vaccination uptake especially in teenagers. In this study, we conducted a population-based study using data from Hong Kong, China to determine the rates of myopericarditis after COVID-19 infection and COVID-19 vaccination, comparing them to background rates and addressing the relative risk of vaccine vs COVID-19 myopericarditis.

## Methods

This population-based retrospective cohort study was approved by the Institutional Review Board of the University of Hong Kong/Hospital Authority Hong Kong West Cluster (UW 20-250). The need for informed consent was waived by the Ethics Committee owing to its observational retrospective nature. Patients who tested positive for COVID-19 by real-time polymerase chain reaction (RT-PCR) at any of the Hong Kong public hospitals or outpatient clinics between 1^st^ January 2020 to 30^th^ June 2021 were included. The data were obtained from the local electronic healthcare database, Clinical Data Analysis and Reporting System, as reported previously ^3^. The primary outcome was myopericarditis. The vaccination data of other countries were extracted using keywords labelled “Myocarditis” and “Pericarditis” upon searching PubMed and the official reports of Hong Kong and the United Kingdom. The data in Hong Kong and the United Kingdom was up to 31^st^ August 2021 and 29^th^ September 2021, respectively.

The rates of myocarditis and pericarditis after COVID-19 infection were calculated by dividing the number of patients by the number of RT-PCR positive COVID-19 patients. The number of doses was defined as the sum of the number of people who received the first and second doses. The hybrid Wilson/Brown method was used to calculate 95% confidence intervals for proportions. The analysis was conducted using PRISM (Version: 9.0.0).

## Results

A total of 11441 COVID-19 patients from Hong Kong were included, of which four suffered from myopericarditis (rate per million: 350; 95% CI: 140-900). This rate was higher than the pre-pandemic background rate in 2020 (rate per million: 61, 95% CI: 55-67) with a rate ratio of 5.73 (95% CI: 2.23-14.73; **Table 1)**. Compared to background rates, the rate of myopericarditis among vaccinated subjects in Hong Kong was substantially lower (rate per million: 8.6; 95% CI: 6.4-11.6) with a rate ratio of 0.14 (95% CI: 0.10-0.19). The rates of myomyocarditis after vaccination in Hong Kong are comparable to those vaccinated in the United States, Israel, and the United Kingdom **(Figure 1)**.

**Table 1.** Incidence rate and rate ratio of myocarditis and pericarditis after COVID-19 infection and vaccination. * Cases for >=12 years old only.

| Outcomes | Background<br>(Hong Kong, China)<sup>8</sup> | Covid-19 infection<br>(Hong Kong, China) | COVID-19 vaccine<br>(Hong Kong, China)<sup>8</sup> | COVID-19 vaccines<br>(US)<sup>9</sup> | COVID-19 vaccines<br>(Israel)<sup>2</sup> | COVID-19 vaccines<br>(UK)<sup>10</sup> |
| --- | --- | --- | --- | --- | --- | --- |
| Diagnosis | Myopericarditis | Myopericarditis | Myopericarditis | Myopericarditis | Myocarditis | Myopericarditis |
| Type of vaccines | NA | NA | CoronaVac<br>BNT162b2 | BNT162b2<br>mRNA-1273<br>Ad26.COV2.S | BNT162b2 | BNT162b2<br>mRNA-1273<br>ChAdOx1 |
| Cases | 416 | 4 | 42 | 37 | 136 | 947 |
| Unit | Persons | Persons | Doses | Doses | Doses | Doses |
| Persons or doses<br>received | 6819672 | 11441 | 4883721 | 3530507 | 10568331 | 93600000 |
| Rate per million<br>persons or doses (95%<br>CI) | 61<br>(55, 67) | 350<br>(140, 900) | 8.6<br>(6.4, 11.6) | 10.5<br>(7.6, 14.4) | 12.9<br>(10.9, 15.2) | 10.1<br>9.5, 10.8 |
| Rate Ratio (95% CI) | 1 (baseline) | 5.73 (2.23, 14.73) | 0.14 (0.10, 0.19) | 0.17 (0.12, 0.24) | 0.21 (0.18, 0.25) | 0.17 (0.16, 0.18) |

**Figure 1.**
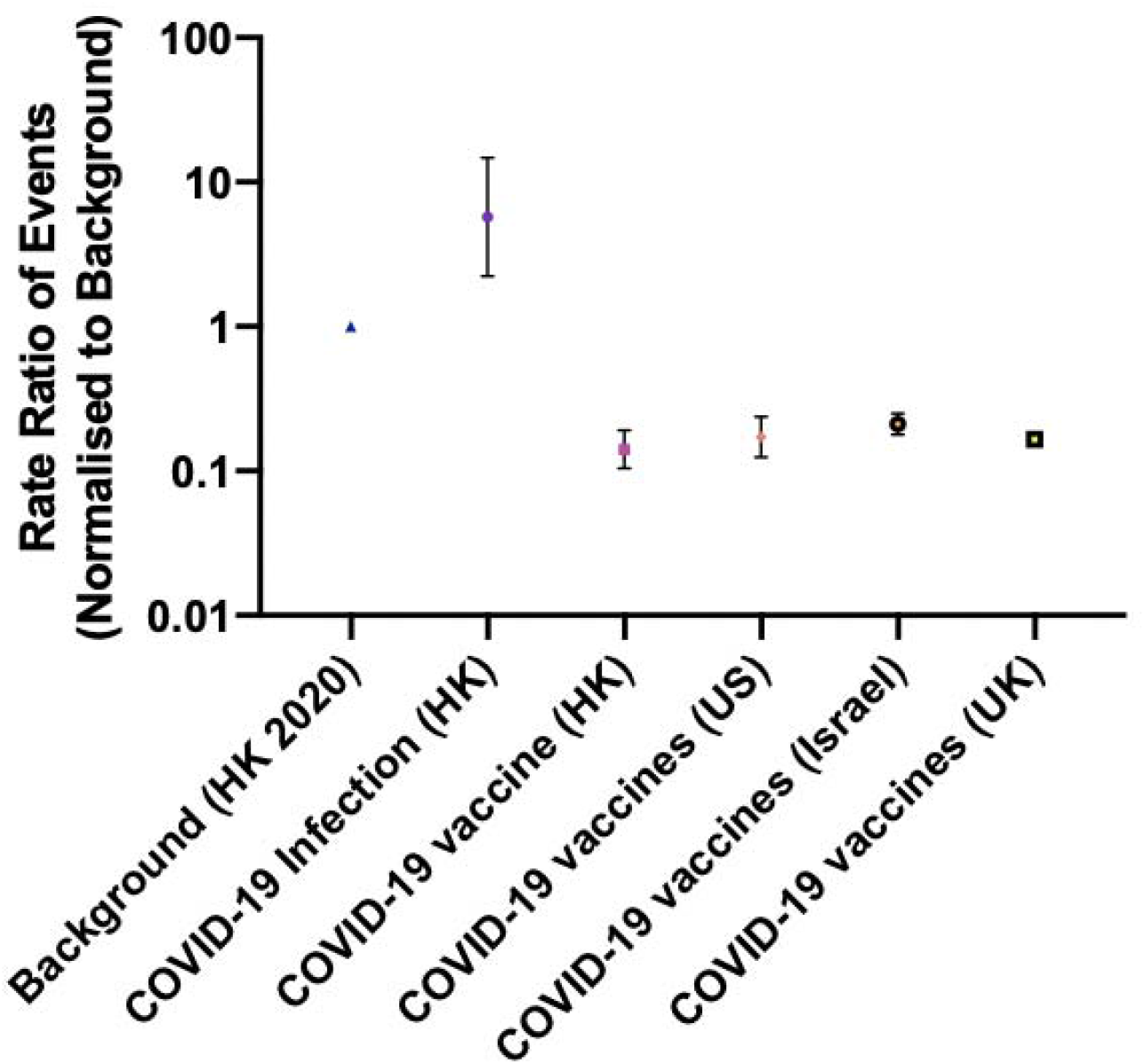
The rate ratio of the events after COVID-19 infection and COVID-19 vaccination.

## Discussion

The main finding from our study is that COVID-19 patients have a higher rate of myopericarditis compared to the pre-COVID-19 era. With COVID-19 vaccination, the rate of myopericarditis was significantly lower than the background rates and the rate in COVID-19 positive patients.

Our team recently developed a predictive model to identify COVID-19 patients at risk of severe disease ^4^. Using an updated dataset, we found that 0.035% of the COVID-19 infected patients developed myopericarditis. This is significantly higher than the baseline, reflecting the association between COVID-19 infection and myopericarditis. Previously, among 718,365 patients with COVID-19, approximately 6.5% developed new-onset myopericarditis.^5^ This higher rate might be an overestimate since most of the patients presented with mild COVID-19 symptoms may not get admitted and diagnosed, thus, not registered with the electronic medical records. Furthermore, our cases represented population-based data locally in Hong Kong, which has practised meticulous contact tracing since the pandemic; the number of total COVID-19 patients in our cohort would, therefore, likely reflect most of the infected patients in the community.

The link between COVID-19 vaccination and myopericarditis has been raised in Israel in May 2021. The results demonstrated that COVID-19 vaccinations are associated with a lower myopericarditis rate than the COVID-19 infected patients. This indicated that the COVID-19 vaccination might protect the vaccinated people from the myocardial injury caused by SARS-CoV-2 infection.^6^ The differences in the outcome suggested that the myopericarditis upon vaccination is unlikely due to the mimicry between the spike protein.^7^ While the rate of myopericarditis is even lower than the background in our data; this does not rule out the causation between the COVID-19 vaccine and myopericarditis. Indeed, the majority of the myopericarditis developed after the second dose of the vaccine among the younger male (<19 years old).^2^ While the rate of myopericarditis following COVID-19 infection is relatively low; further studies are needed to characterise the mechanism of myopericarditis after COVID-19 vaccination. Nonetheless, with the current increasing numbers in COVID-19 globally, it would appear that ever person will come in contact with SARS-CoV-2, therefore the overall risk of myopericarditis will be much higher at a population level without vaccination.

### Strengths and limitations

This study has several strengths. Firstly, COVID-19 cases were identified by RT-PCR testing across the public sector. Therefore, missing cases are likely to be few. Secondly, possible cases of vaccine-related myopericarditis were reviewed by an expert panel, that examined the medical records independently. This adjudication has permitted the accurate classification of cases according to established international guidelines. Nevertheless, had we included possible cases rather than cases definitely linked to vaccinations, the rate ratios of myopericarditis cases in infected patients to cases occurring after COVID-19 vaccination would be even lower, and therefore does not alter our conclusion.

However, several limitations should be noted. Firstly, the cohort included patients recruited from a single region and as such, is unable to account for any geographical heterogeneity that may exist. Secondly, the vaccination data reported in the local Department of Health did not provide the number of myopericarditis after the first and the second dose of vaccination and therefore the results are not stratified. Thirdly, we might have missed COVID-19 myopericarditis in patients who might have died from acute heart failure complications due to myopericarditis, but recorded as heart failure deaths. This however, would have increased the difference between COVID-19 and vaccine related myopericarditis, further strengthening the usefulness of vaccination.

## Conclusion

COVID-19 infection is associated with significantly higher rate of myopericarditis compared to both background rates and vaccine associated myopericarditis.

## Data Availability

All data produced in the present study are available upon reasonable request to the authors

## Central illustration

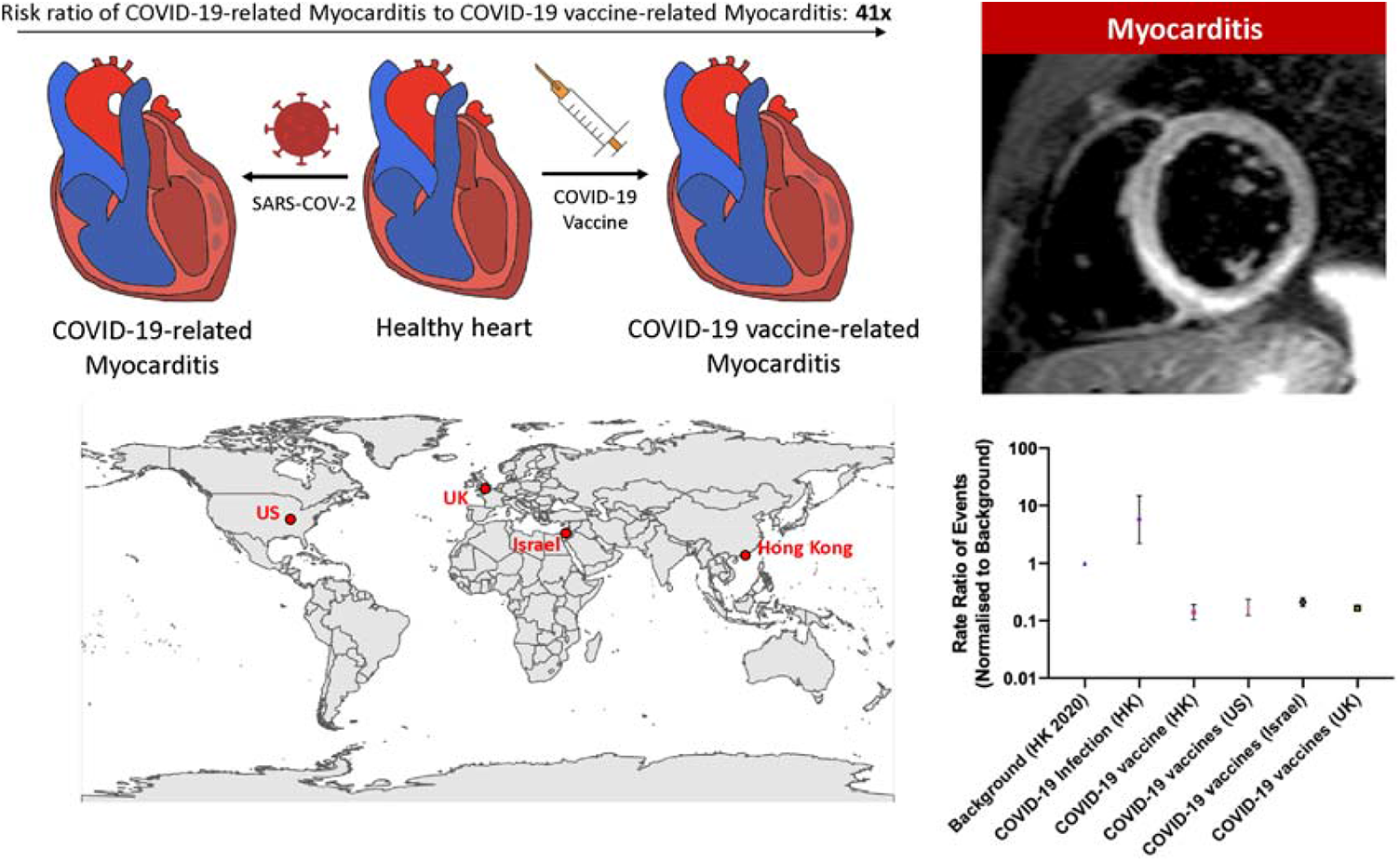

## Notes

### Competing Interest Statement

The authors have declared no competing interest.

### Funding Statement

This study did not receive any funding

### Author Declarations

This population-based retrospective cohort study was approved by the Institutional Review Board of the University of Hong Kong/Hospital Authority Hong Kong West Cluster (UW 20-250).

